# Metabolic Physiological Networks: The Impact of Age

**DOI:** 10.1101/2020.08.05.20168997

**Authors:** Antonio Barajas-Martínez, Jonathan F. Easton, Ana Leonor Rivera, Ricardo Martínez-Tapia, Lizbeth de la Cruz, Adriana Robles Cabrera, Christopher R. Stephens

## Abstract

Metabolic homeostasis emerges from the interplay between several feedback systems that regulate the physiological variables related to energy expenditure and energy availability, maintaining them within a certain range. Although it is well known how each individual physiological system functions, there is little research focused on how the integration and adjustment of multiple systems results in the generation of metabolic health. The aim here was to generate an integrative model of metabolism, seen as a physiological network, and study how it changes across the human lifespan. We used data from a transverse, community-based study of an ethnically and educationally diverse sample of 2572 adults. Each participant answered an extensive questionnaire and underwent anthropometric measurements (height, weight, waist), fasting blood tests (glucose, HbA1c, basal insulin, cholesterol HDL, LDL, triglycerides, uric acid, urea, creatinine), along with vital signs (axillar temperature, systolic and diastolic blood pressure). The sample was divided into 6 groups of increasing age, beginning with less than 25 years and increasing by decades up to more than 65 years. In order to model metabolic homeostasis as a network, we used these 15 physiological variables as nodes and modeled the links between them, either as a continuous association of those variables, or as a dichotomic association of their corresponding pathological states. Weight and overweight emerged as the most influential nodes in both types of networks, while high betweenness parameters, such as triglycerides, uric acid and insulin, were shown to act as gatekeepers between the affected physiological systems. As age increases, the loss of metabolic homeostasis is revealed by changes in the network’s topology that reflect changes in the system-wide interactions that, in turn, expose underlying health stages. Hence, specific structural properties of the network, such as weighted transitivity, can provide topology-based indicators of health that assess the whole state of the system.

## 1. Introduction

Metabolic homeostasis arises from the interchanges between multiple chains of biochemical reactions and their mechanical responses. These exchanges maintain variables related to energy expenditure and energy availability within suitable ranges for the organism. The components of these chains are shared by multiple others, thereby constituting a metabolic network. Unfortunately, many processes of this network are not readily accessible in the clinical setting. Therefore, to make inferences about the underlying energy metabolism, various biomarkers-either biochemical or anthropometric-have been used to assess the state of the different physiological sub-systems that constitute the network. These physiological variables represent either regulated variables or physiological response systems (Fossion, Rivera, & Estañol, 2018). The lability of the values of physiological response variables, and the consequent stability of regulated variables, characterizes the robustness of a complex homeostatic system that resorts to pathological states only in order to preserve vital variables (Kitano et al., 2004). Thus, homeostasis can be established by the interplay between physiological variables, allowing its study through a metabolic physiological network.

Over time, the physiological compensatory systems that maintain homeostasis become worn down due to the cumulative impact of metabolic insults, transitioning from healthy to maladaptive states that precede disease onset (Stephens et al., 2020). An already existing medical notion of this system-wide progression of states before the overt onset of disease is metabolic syndrome (MetS), whose prevalence increases strongly with age (Hildrum et al., 2007) and unhealthy lifestyles. At early stages, MetS biomarkers indicate invisible alterations, wherein homeostasis can still be preserved (Huang, 2009). Insulin resistance, dyslipidemia, endothelial dysfunction, prothrombotic, proinflammatory states and, more recently, oxidative stress are then employed to diagnose a condition of increased cardiometabolic risk (Reaven, 1993; Vona et al., 2019). With this in mind, several medical organizations established operational diagnostic criteria (Xu et al., 2018), starting with preexisting diagnostic thresholds for each associated disease, and then lowering them in order to provide a preventive focus for the diagnosis of MetS (Parikh & Mohan, 2012). In the continued presence of metabolic insults, as each physiological regulatory system fails, the cascade is absorbed downstream by the next system. Eventually, what were originally reversible pathological states progress to become irreversible diseases. This is the final stage, characterized by the lability of the regulated variables, wherein the physiological response systems become overwhelmed. These states correspond to clinical diseases that were the basis for the first historical descriptions of MetS, where gross anatomical changes and clinically overt symptoms, comprising obesity, hypertension, gout, atherosclerosis and obstructive apnea were first associated (Enzi et al., 2003). However, it is usually on a scale of decades that these physiological interactions change substantially. Disease appears only once the robustness of the metabolic physiological network is broken, and regulated variables lose their tight control.

The current approach to determining metabolic health relies on using the thresholds of individual biomarkers, without considering the overall physiological network itself. As threshold values are the result of a compromise between sensitivity and specificity, they must be tailored adequately for both screening and diagnostic purposes in each population (Almeda-Valdes, et al., 2016). However, current thresholds consider neither age stratification nor the duration of the pathological states, resulting in medical interventions that are targeted towards single variables and only late in life (Easton et al., 2019). Furthermore, standard of care for these complex states is no different from the treatment of each of its individual components (Kahn, 2007). Although targeted approaches for age have been proposed, for providing further insight on the etiology of risk factors and guide disease-prevention strategies (Xu et al., 2019; Leatherdale, 2015; Leventhal, Huh, & Dunton, 2014), it has been argued that the principle utility of MetS as a concept relies on the preventive nature of its scope, and the idea that single interventions could improve simultaneously all of the current five MetS criteria (Vassallo, Driver, & Stone, 2016). However, there is still doubt as to how to weight the risk associated with each factor, or their combinations (Sattar, 2008). Indeed, given the increasing abundance of metabolic biomarkers that predict disease, there is not even a universal consensus on which criteria should be included and excluded in the first place in order to best assess metabolic health (O’Neill & O’Driscoll, 2015). As metabolic health is an emergent property, arising from the interaction of multiple physiological systems over time, the framework of complexity provides the means for a whole-system analysis (Haring et al., 2012; Lusis, Attie, & Reue, 2008; Sun et al., 2012), rather than a reductionist variable-by-variable approach. In previous work (Stephens et al., 2020), we considered how ageing was an important driver of metabolic change across a wide variety of metabolic biomarkers (anthropometric, fasting blood test and vital signs measurements), considering each one individually and noting a substantial degree of heterogeneity as to the impact of aging across them. In contrast, in the present study, we have used networks of these biomarkers as a means to give a more holistic, systems-biology perspective in order to demonstrate how the changes in the coupling between regulated variables and those regulatory systems that try to maintain homeostasis lead to metabolic health changes over a lifetime. In particular, in this paper, we will use complex physiological networks to better understand these interactions, constructing a data-driven network of biomarkers that can be used to characterize homeostasis and how it changes as a function of age.

## 2. Results

### 2.1 Demographic description of the population

A general description of our study population (n=2572), and the distinct age groups is provided in Table 1. The mean age of the participants was 38 years old (standard deviation, SD= 15, range from 18 to 81 years old). Our population sample was predominantly female (65%). This predominance was preserved across all of the age groups considered with no statistically significant differences between groups. Our population sample comes mainly from the metropolitan region of Mexico City (93%), with the remaining participants from neighboring states. Educational level proportions changed within the age groups, with an increasing trend for postgraduate and basic education (at most 12 years of study), and a decreasing trend for undergraduate education, that are illustrative of the population composition within the sample (Table 1). We found that MetS prevalence, as defined by the criteria in (Alberti et al., 2009), increased significantly by age (under a chi-squared test for trend p<0.001), beginning with a prevalence of 4% for the first age group (<25 years old), which increased ten-fold to 47% in the age group from 55 to 65 years old. For adults older than 65 years old, MetS prevalence is high (43%) but is lower than that from 55 to 65, however, this difference between groups is not statistically significant *(X^2^* (1, N = 659) = 0.14, p = 0.7).

**Table 1.**
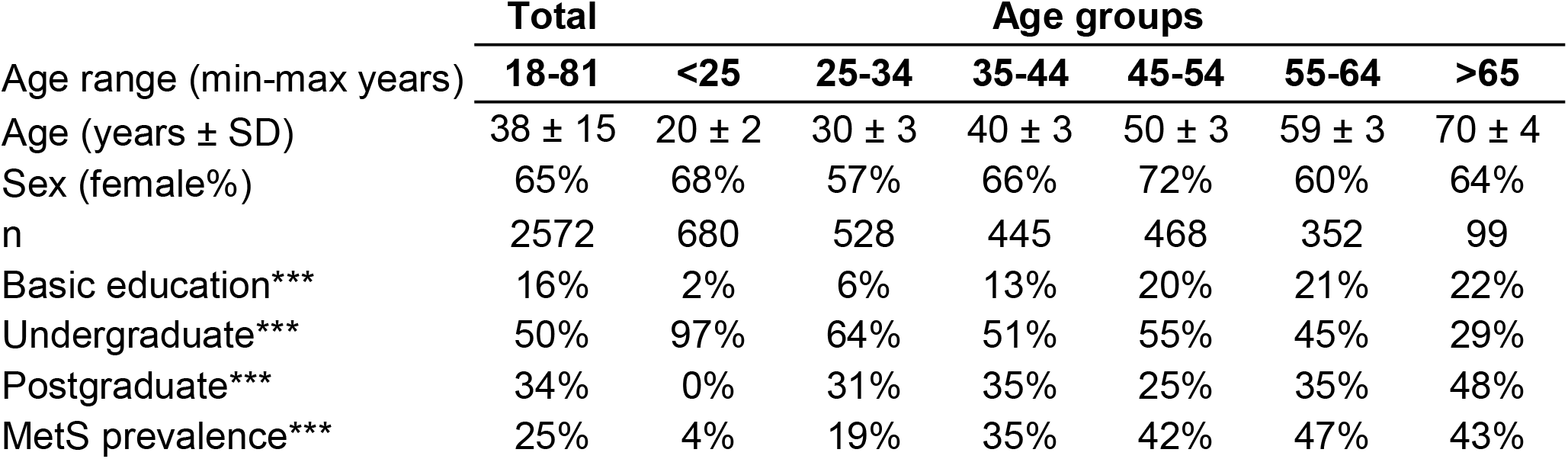
Demographic description of the population. General description of the total sample and the age groups is provided with sex (female percentage), mean age ± SD and total number of participants in each group. The presence of a trend with age by chi-squared tests for trends is indicated by *** for p<0.001.

### 2.2 Physiological variables and pathological state prevalence change with age

To examine whether this increase in MetS prevalence with age was due to an increment in the mean values of the physiological variables or to an increase in the tail of the distribution above the cut-off values (Table 2), linear regressions and chi-squared tests for trends were evaluated (Table 3). Most of the physiological variables (fasting glucose, HbA1c, LDL cholesterol, triglycerides, urea, creatinine, waist, weight, systolic and diastolic blood pressure) increased progressively with age, having a statistically significant positive linear regression slope, whereas height and axillar temperature decreased, being associated with a statistically significant negative linear regression slope. In contrast, three physiological variables: basal insulin, HDL cholesterol, and uric acid, showed no linear changes as a function of age. Following the trend of their respective physiological variables, the prevalence of pathological states also grew with age, with one exception: high temperature. While changes in the mean values of the physiological variables as a function of age were considerably smaller, as shown by the slopes in the linear regressions, the proportion of the population above the cut-off values for the pathological states increased substantially (Table 3). For the physiological variables, waist circumference, weight, systolic and diastolic pressure had the greatest regression coefficients as a function of age. Regarding the prevalence of pathological states, overweight, low estimated glomerular filtration rate (eGFR), and hyperglycemia, had the greatest increase as a function of age, followed by high blood pressure, high LDL, hypertriglyceridemia, high HbA1c, and azotemia. Age had a widespread influence on most of the components of MetS, whether regarded as continuous or as categorical variables. The prevalence of low HDL and hyperuricemia changed with age, although this trend was not detected by a linear regression.

**Table 2.**
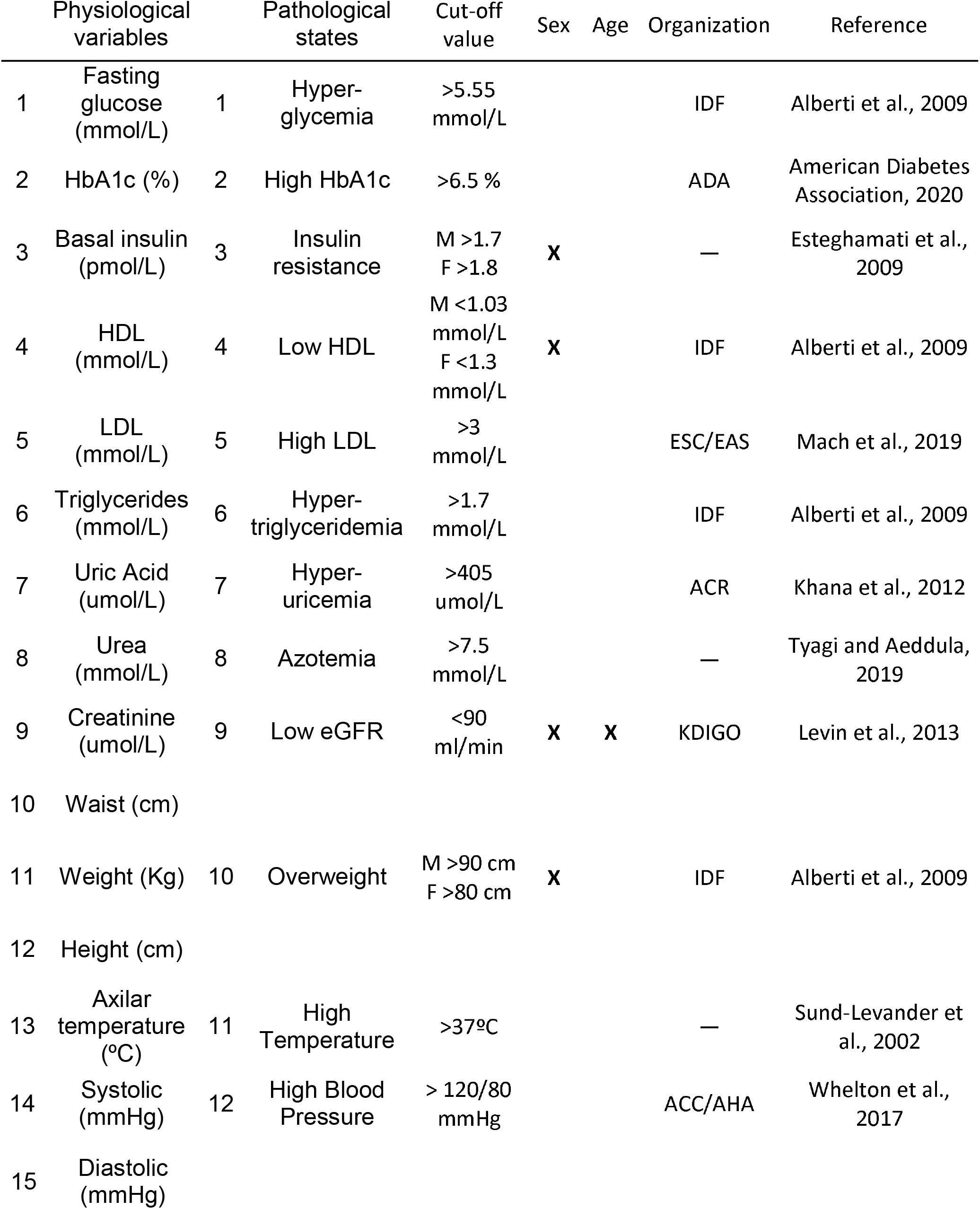
Pathological states criteria. Threshold values employed for the classification of pathological states. Current criteria that are tailored for age and sex are indicated in the columns.

**Table 3.**
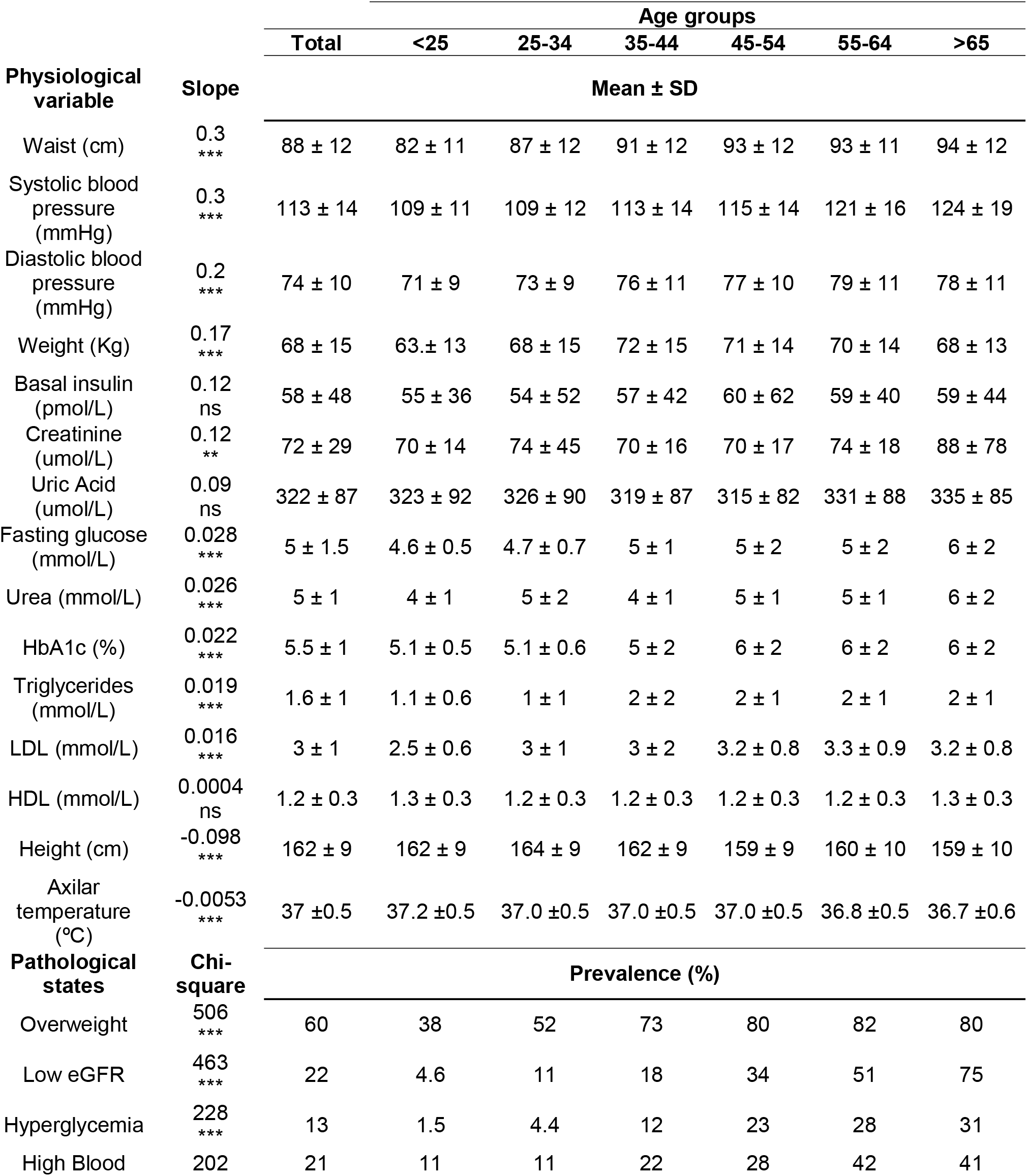
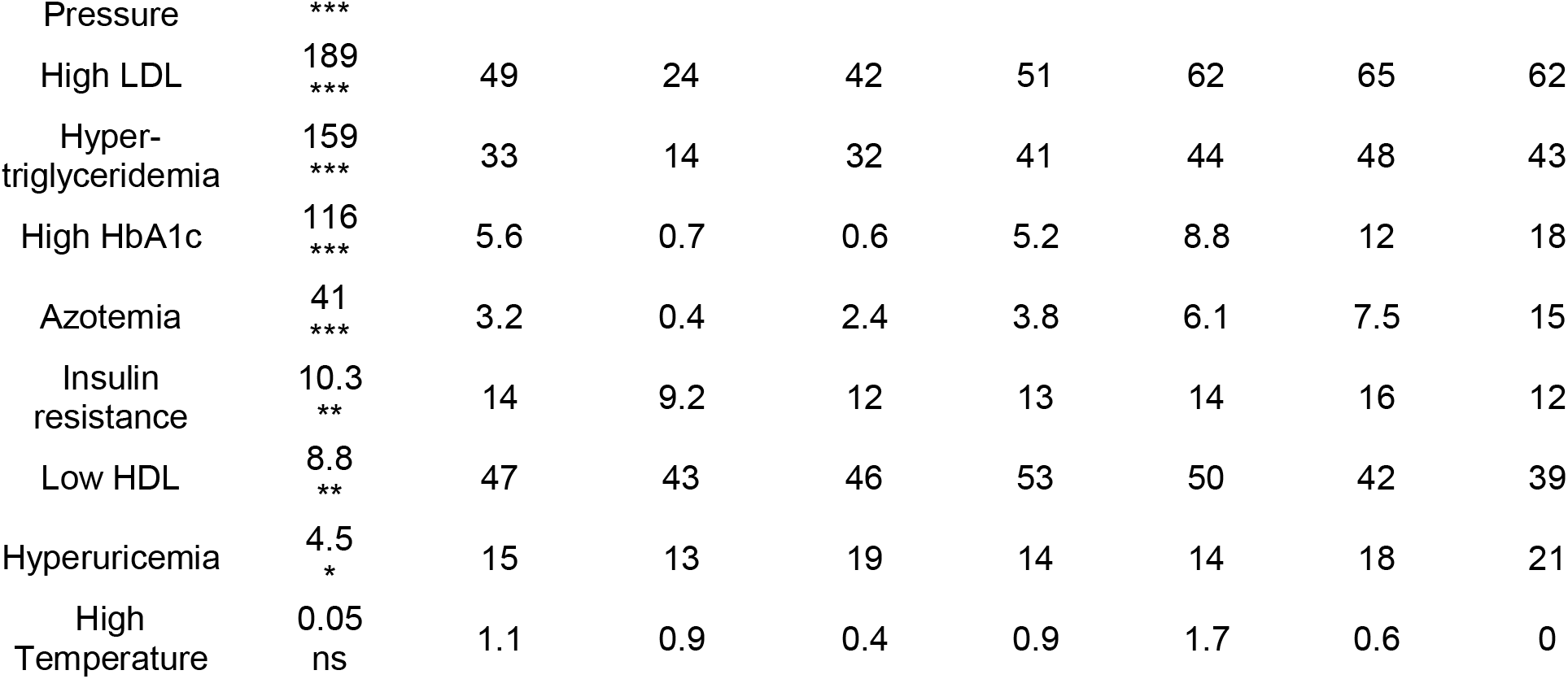
Physiological variables means and pathological states prevalence. Mean ± SD of each of the 15 physiological variables and prevalence of each of the 12 pathological states are displayed for each group. Linear regressions for the physiological variables, and chi-squared tests for trends of the pathological states prevalence are given. Significance of the trend with age is indicated by *for p<0.05, ** for p<0.01, and *** for p<0.001.

### 2.3 Metabolic modules can be identified within the network

To investigate how metabolic physiological components are grouped within the networks, we employed two strategies, either identifying largest cliques or finding clusters within the networks (see Figure 1). For the first strategy, the largest cliques method shows the biggest possible, maximally connected subgraphs of a network, indicating which components go hand in hand most frequently across distinct age groups (Figure 1C and 1D). For the physiological network, weight, waist circumference, uric acid, systolic and diastolic blood pressures appeared most frequently in the major cliques (Figure 1C). In the pathological states network, insulin resistance, hypertriglyceridemia, overweight and hyperglycemia were most frequently found to occur within the largest cliques (Figure 1D). For the second strategy, the networks were assorted into different clusters, using the Louvain algorithm (Blondel et al., 2008) for the physiological network, or the Spinglass algorithm (Reichardt & Bornholdt, 2006) for the pathological states network (Figure 1E and 1F). Four main clusters were found in the physiological network (Figure 1E), with the main cluster associated with weight, followed by a cluster around urea. An intermediary cluster was found around glucose and HbA1c, while systolic and diastolic blood pressure remained separated from the rest. For the pathological states network, the main cluster was around hyperglycemia and the second was around low eGFR, with an intermediate cluster around high blood pressure and high temperature (Figure 1F). The metabolic components within these clusters were related by metabolic pathways, establishing metabolic modules.

**Figure 1.**
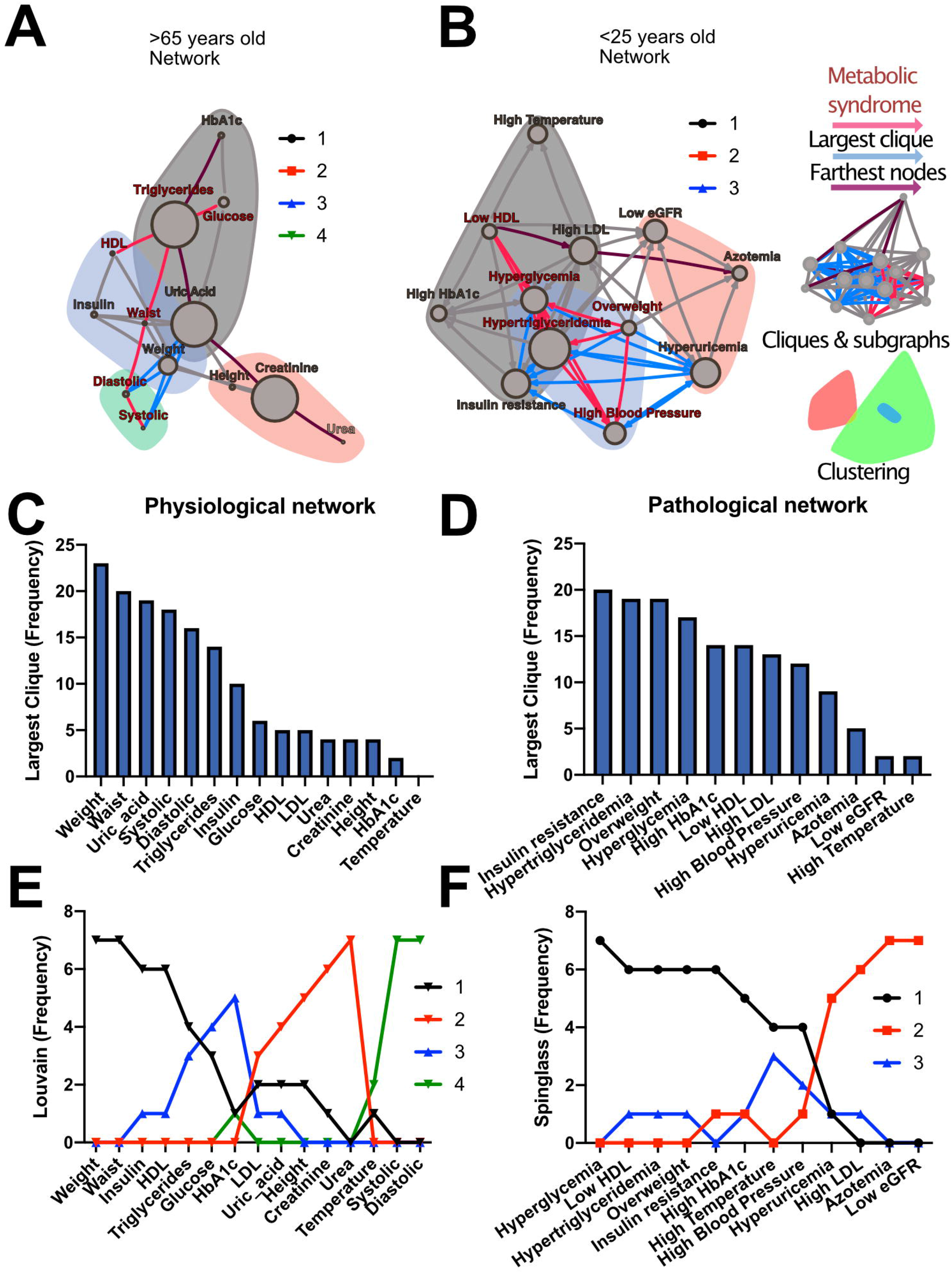
Physiological subsystems identified by Data-driven association. Representative networks (A) for the physiological variables network and (B) for the pathological states network. Physiological variables and pathological states clusters are shown as largest cliques (blue connections), and, as clusters (nodes within color highlighted areas). In both metabolic physiological networks, the red subgraph shows the currently accepted MetS components. The diameter of the network - the two furthest nodes path - is highlighted in purple. (C) Frequency of physiological variables composing the largest clique of each age group network. (D) Frequency of pathological states fully associated within largest cliques as shown by the pathological states network. The frequency of appearance of a node pertaining to a certain cluster (membership) was registered. Since 7 networks were generated (all participants, and 6 age-range groups), a node belonging to the same cluster across the entire lifespan would reach a value of 7. In (E), the frequency value represents how many times a node is part of the same cluster for the physiological variables, where the Louvain algorithm was used to determine clusters. Three main clusters appear, with blood pressure variables making a fourth. (F) Cluster membership of pathological states using the spinglass community algorithm that selects the group of nodes most likely to be found in the same state. Three main clusters appear, with different groups of pathological states in each one.

Both strategies lead to a selection of nodes that differs from current MetS criteria (Figure 1A and 1B). While waist and weight are frequently part of the largest clique of the network, they are often clustered separately from the metabolic components of triglycerides and glucose. Triglycerides, both as a physiological variable or as pathological state, are frequently part of the largest cliques and belong to the main cluster of the networks. Hyperglycemia, on the other hand, is part of the main cluster only in the pathological states network and is frequently part of largest cliques but is not part of the largest cliques nor of the main cluster as a physiological variable (glucose). Systolic and diastolic blood pressures are also frequently part of the largest cliques, but only as physiological variables and not as a pathological state. They belong mainly to the cluster of overweight as pathological states, but are in an independent cluster as physiological variables. Finally, HDL cholesterol as a physiological variable was seldom part of the largest cliques, however, it was part of the main cluster in the pathological states network.

### 2.4 The role of metabolic biomarkers within the network across a lifetime

The relations between the physiological variables and pathological states within the networks change with age. We observed that obesity, whether as proxied by the weight and waist circumference physiological variables, or as the overweight pathological state, is the main influencer in the network as measured by eigencentrality, a role which remained stable across all age groups (Figure 2A and 2C). In contrast, physiological variables with characteristically tight homeostatic control, like glycemic variables and temperature, were uninfluential in the network (Figure 2C). For the pathological states network, the largest influence, as measured by hubscore (a generalization of eigencentrality for directed graphs), was exerted by overweight, with the components of dyslipidemia becoming less influential from 25 to 34 years old onwards, while the pathological states associated with low estimated glomerular filtration rate (low eGFR) steadily became more relevant above 65 years old (Figure 2B and 2D). Gatekeeping biomarkers of the flow between systems were uric acid, insulin, HbA1c and HDL in the physiological network, while hypertriglyceridemia, insulin resistance, hyperglycemia and high HbA1c were the main intermediaries between pathological states (Figure 2E and 2F). Unlike eigencentrality values, flow betweenness values change profoundly as a function of age (Figure 2E and 2F).

**Figure 2.**
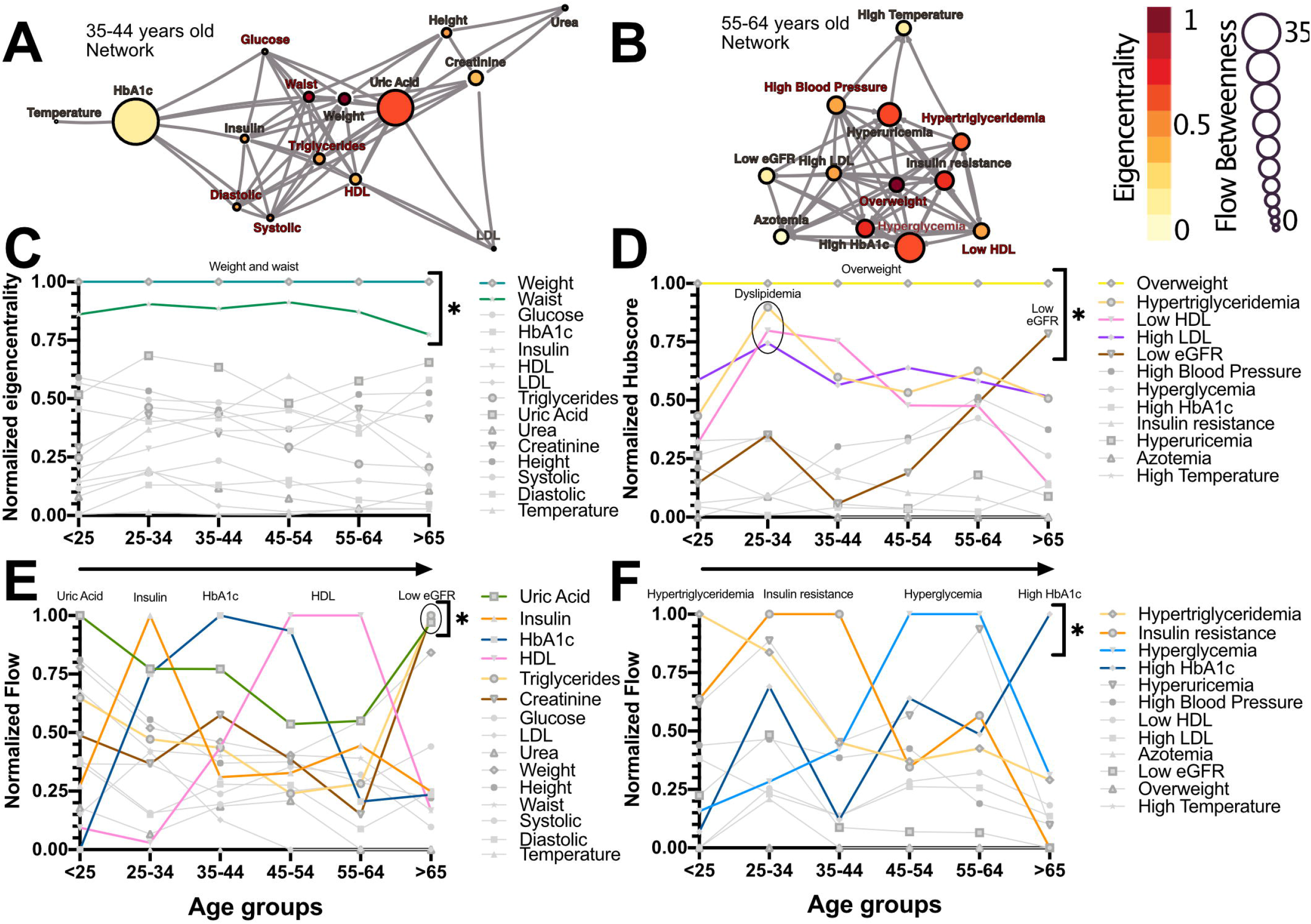
Network modeling highlights physiological and pathological interactions. Centrality measurements identify the role of each physiological variable or pathological state within the metabolic network. (A) Physiological network from 35 to 44 years old, and (B) pathological network from 55 to 64 years old, as examples of the different centrality contribution that each node has. Influence is measured by eigencentrality and is represented by node color, while betweenness is measured by flow and represented by node size. The values from these examples are emphasized inside gray rectangles. (C) Most influential nodes in the physiological variables network, Weight and waist, are indicated. (D) Most influential nodes as seen by eigencentrality in the pathological states network. Overweight, dyslipidemia and low eGFR are indicated. (E) Gatekeeping nodes, as seen by flow betweenness, that mediate the associations between those physiological variables that are not directly connected. (F) Gatekeeping nodes that are the route between unconnected pathological states. The most meaningful nodes in this regard are hypertriglyceridemia, insulin resistance, hyperglycemia and high HbAlc as age increases. 0 indicates values unlikely to be found by chance alone in CUG tests.

### 2.5 Whole network topology as a biomarker for metabolic homeostasis

As well as local properties of the physiological variables and pathological states networks, global properties also change with age. Topological properties of these networks for all the age groups are summarized in Table 4. Noticeably, for the pathological states network, we found that reciprocity was lower than would be expected, while transitivity of the networks was greater than that expected for comparable networks of the same size, number of links or dyads (Table 4). Characteristic path length was lower than would be expected for random networks. Moreover, the local transitivity of physiological variables reaches a peak in the life decade between 25 and 34 years old, and from then on, the transitivity begins to decrease (Figure 3A and 3C). However, this decrease is not the result of a reduction in the weighted degree distribution (strength) of the correlations within the network, which are similar across all age groups (Figure 3E), instead it is related to an increase in the number of edges within the network, as presented by network density (Table 4). In other words, the organization of the physiological variables changed independently from the strength of the relationships between the variables. Over a lifetime, nodes within a cluster tend to connect more within themselves rather than outside the cluster. This topological change results in a modularity increase in the physiological network (Figure 3D). However, this trend was not shared with the pathological states network. In this network, there is a trend towards increasing transitivity until the 45 to 54 years old age groups group, and a decrease in older groups (Figure 3B and 3C). Pathological states became increasingly correlated as a function of age, until reaching a maximum in the decade between 45 and 54 years old (Figure 3C). This clustering change is related to the weighted degree distribution of the pathological states network (Figure 3F) and to an increase in the density of the network (Table 4). In these networks, modularity decreases from the 35 to 44 years old group onwards (Figure 3D). Three stages become apparent: a healthy stage, where the clustering of both networks increases; a transition stage, where the clustering of pathological states increases, while the clustering of physiological variable decreases; and a disease stage, where the clustering of both networks decreases (Figure 3C). The proportion between clustering coefficient and characteristic path length in a network can be summarized by the small world index to compare structural changes in our matching networks of increasing age. For the physiological networks of groups starting below 54 years, the small-world index has values between 1.3 and 1.9, increasing to values above 2 in the groups above 55 years old. All pathological networks had a greater small world index than the corresponding physiological networks, which increased substantially in the age group above 65 years old and concurrently with a decrease in the global clustering coefficient.

**Figure 3.**
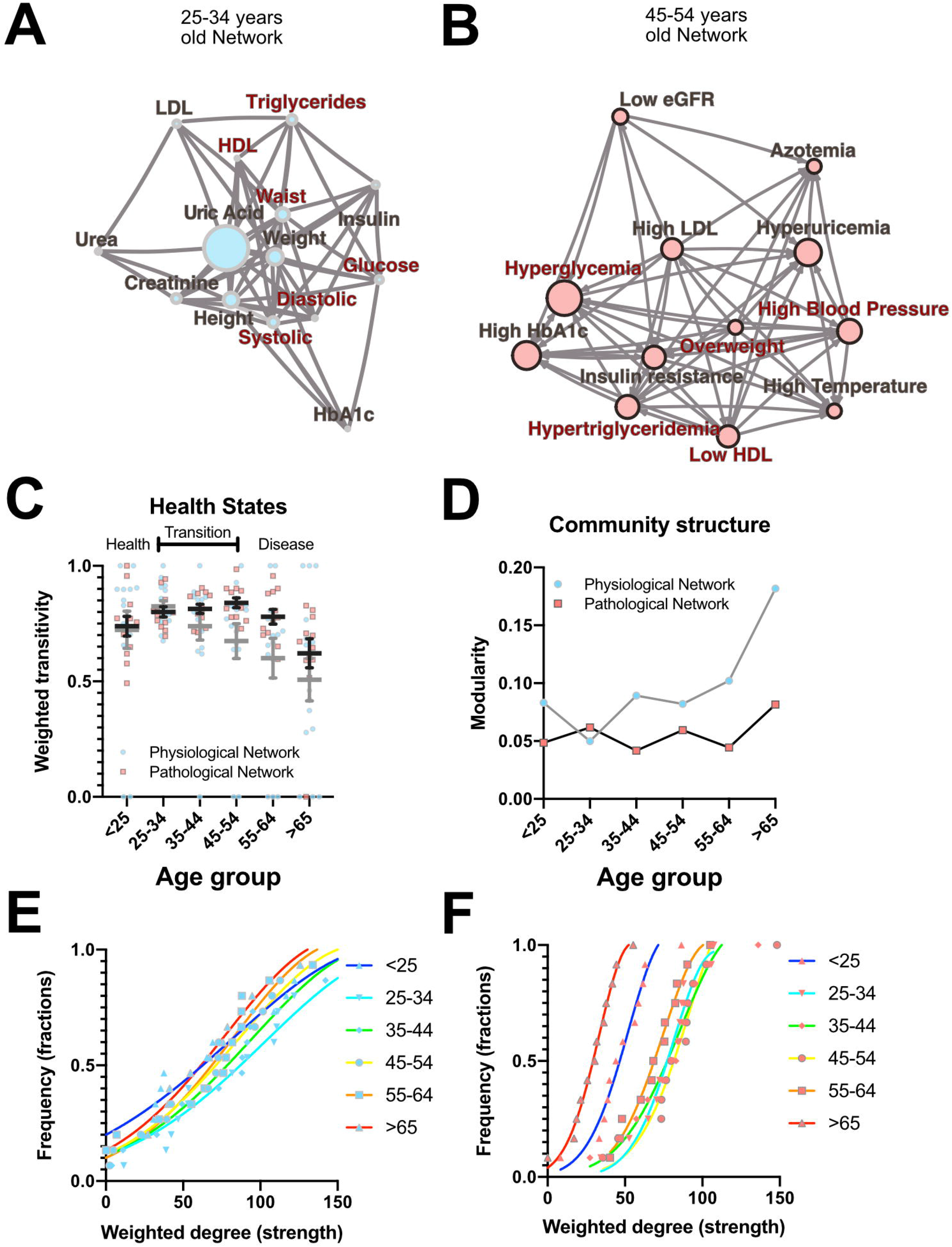
Topological properties from physiological and pathological networks. Network structural changes as a function of age can be seen using several topological metrics. (A) Physiological network of the third decade of life as a visual example of weighted transitivity in a tightly intertwined network. (B) Pathological network of the fifth decade of life as an example of weighted transitivity in a directed network. These two networks represent the greatest transitivity in all age groups. (C) Weighted transitivity of each network as the mean ± S.E.M. from all life decades, n=2572. The values that come from the physiological network nodes are highlighted in blue and for the pathological states network in pink. (D) Weighted transitivity of each network as the mean ± S.E.M. value of the 12 tested pathological states from all the age groups. Frequency distribution of the weighted degree (strength) of the network in each life decade (E) for the physiological networks and (F) for the pathological states networks. Age dissociates physiological variables, as seen by the reduction of the weighted transitivity in the physiological network, but without a significant change in the weighted degree, while pathological conditions become more associated with age, as seen in the pathological network, reaching a peak at the fifth decade of life.

**Table 4.** Topological properties of the physiological variables and pathological states networks. Global measurements for topological properties are shown for each network. In the case of the pathological states network, since it is directed, reciprocity of the network is also shown.

|  | Total | Age groups |  |  |  |  |  |
| --- | --- | --- | --- | --- | --- | --- | --- |
|  |  | <25 | 25-34 | 35-44 | 45-54 | 55-64 | >65 |
| Physiological variables networks |  |  |  |  |  |  |  |
| Density | 0.73 | 0.47 | 0.63 | 0.53 | 0.49 | 0.39 | 0.26 |
| Global transitivity | 0.79 | 0.71 | 0.74 | 0.70 | 0.70 | 0.67 | 0.47 |
| Characteristic path length $L$ | 1.27 | 1.38 | 1.40 | 1.57 | 1.67 | 1.54 | 1.85 |
| Clustering coefficient $C$ | 0.84 | 0.67 | 0.78 | 0.69 | 0.63 | 0.58 | 0.51 |
| Smallworld Index | 1.3 | 1.9 | 1.3 | 1.4 | 1.5 | 2.1 | 2.3 |
| Pathological states networks |  |  |  |  |  |  |  |
| Density | 0.48 | 0.33 | 0.40 | 0.40 | 0.42 | 0.39 | 0.30 |
| Reciprocity | 0.07 | 0.10 | 0.04 | 0.06 | 0.04 | 0.06 | 0.03 |
| Global transitivity | 0.91 | 0.69 | 0.79 | 0.79 | 0.83 | 0.78 | 0.67 |
| Characteristic path length $L$ | 1.19 | 1.27 | 1.15 | 1.23 | 1.20 | 1.25 | 1.09 |
| Clustering coefficient $C$ | 0.50 | 0.41 | 0.41 | 0.43 | 0.43 | 0.42 | 0.32 |
| Smallworld Index | 2.8 | 3.5 | 3.3 | 3.1 | 3.1 | 3.1 | 4.6 |

## 3 DISCUSSION

Metabolic homeostasis loss is the main driver of noncommunicable diseases and their resulting mortality. These complex diseases involve diverse combinations of biomarkers of risk that occur more often together than by chance alone (Alberti et al., 2009). Currently, however, only five such factors are monitored for the assessment of metabolic health (overweight, high triglycerides, low HDL cholesterol, high systolic blood pressure, and high fasting plasma glucose). By adopting a network approach, in this study, we have shown that, in reality, not only the level of each individual factor is important, but also their correlations, both local and global. Local properties of the network are equivalent to current reductionist approaches, while global properties provide new metrics that can be used as markers of metabolic health. As allostatic load on body metabolism increases with age, changes in the ratios between different physiological variables represent the adaptive adjustment of their corresponding setpoints in order to accommodate an increasing burden of internal failures and cumulative external insults (Fossion et al., 2018; Goldstein, 2019). Here, we have shown that the number of correlations present within the networks, represented as network density (Table 4), the number of connections of each node, represented as the node’s degree, and the strength of the correlation, represented as weighted degree, all change gradually across age groups and reflect this adaptive adjustment (Figure 3). Therefore, topological properties that emerge from the structure of the networks reflect how whole-system interactions within the physiological network change over a lifetime and, in particular, show how, as age increases, the loss of metabolic homeostasis is revealed by these changes. For example, local weighted transitivity measures the probability that the neighbors of a node are connected among themselves. This measure has the advantage of being largely independent from the size of the network (Barabási et al., 2003). Changes in this metric give insight into how the cumulative impact of metabolic insults increases and decreases the relations between physiological variables and pathological states. At the global level, transitivity and the clustering coefficient of the network are two indicators of how the network’s connections become aggregated or disaggregated as a function of age. Therefore, these changes in the networks’ structure echo the underlying homeostatic changes.

The transition from health to disease, in the case of complex diseases, can be described by three-state models (Chen et al., 2017). In the healthy stage, regulated variables are kept within strict bounds and physiological response systems increase their activity proportionally in order to compensate the impact of interaction with the environment. In the transition stage (from 35 to 54 years old) regulated variables increase their correlation with their physiological response system as metabolic insults are not fully compensated. At this stage internal malfunctions can be buffered, but at the expense of the development of pathological states, that then begin to correlate, leading to an ever increasing burden (Figure 3). Finally, homeostasis is lost, and pathological states lead to disease onset in an irreversible fashion, resulting in a decrease in the clustering of both network types. Regulated variables are now fully dysregulated from their corresponding regulatory system variables and correlations are lost. Our results show that the transition from health to disease is reflected in our topological metrics as a result of the changes in the correlations between physiological variables and the corresponding association between pathological states. The different network metrics we evaluated show that our networks are not random (Table 4). Although a formal, large-scale topological characterization of our physiological networks falls beyond the scope of this work, and would potentially require the addition of many more variables, it is interesting to point out that the observed properties of scale free and small world are properties that are frequently found in complex biological systems (Song et al., 2005). It has been argued that these topologies confer properties of network robustness and adaptability that are desirable as properties with a homeostatic interpretation (Fossion et al., 2018; Toledo-Roy, Rivera, & Frank, 2019). Nevertheless, considering the wide structural diversity found in real-world networks, classification of these complex systems remains an active area of development (Broido & Clauset, 2019; Hilgetag & Goulas, 2016).

**Figure 4.**
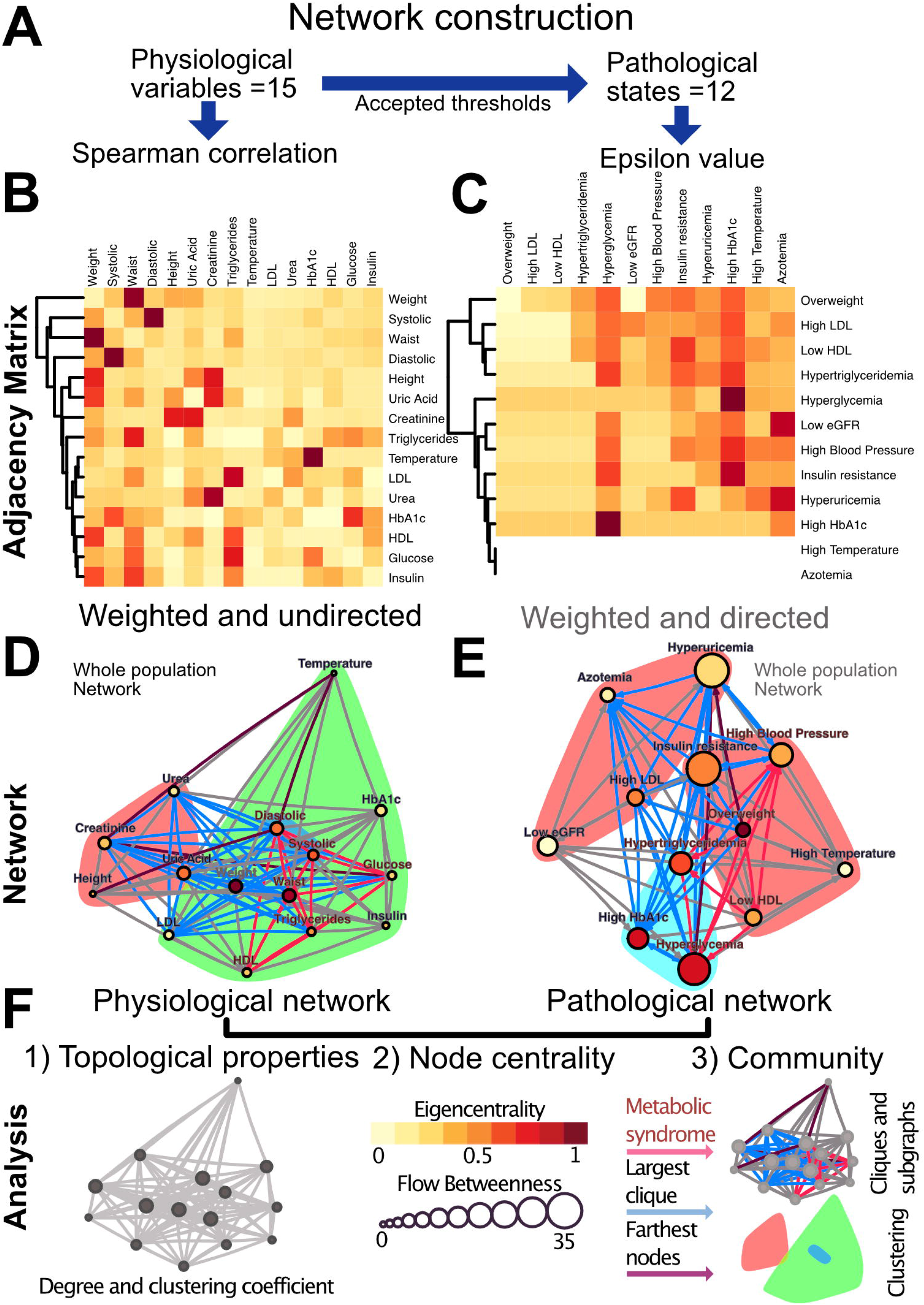
Metabolic physiological network construction from matrices. (A) Correlation of 15 physiological variables and their corresponding 12 pathological states associations were modelled using Spearman correlation and £ value, respectively. (B) Adjacency matrix as a heatmap where the darker the red indicates a greater monotonic relationship between two physiological variables, as calculated by the Spearman rank correlation rho. (C) *ε* value between each pair of pathological states, a darker red indicating a greater probability of coexistence. In both heatmaps, rows and columns are ordered by weighted degree, and on the left side of the heat maps the resulting hierarchical dendrogram is shown. For directed networks some nodes lacked outgoing links, this is presented as blank rows. (D) Undirected network of physiological variables for the whole sample. The edges are weighted by the rho value in the Spearman correlation. The size of the node shows the flow betweenness of a node, the eigencentrality is shown by its colour and the colour shadowed areas indicate the Louvain clusters. (E) Directed network of pathological states. The edges are weighted by the value, the size of the node shows the flow betweenness of each node, the eigencentrality is shown by its colour and the colour shadowed areas indicate spinglass clusters. In both networks, the red subgraph shows the components of MetS, while the blue subgraph highlights the largest clique and the diameter of the network is in purple. For Spearman correlation, values with p>0.001 were discarded, whereas for £, values below 1.96 were discarded.

Individual biomarkers were described in the context of the network through centrality measurements of influence and intermediacy. The important influence of weight on the metabolic network was found in both network approaches and was sustained across all age groups (Figure 2). Additionally, weight-associated physiological variables and their corresponding pathological states were most frequently embedded within the largest cliques. Both these results exhibit the central role of weight inside the metabolic networks. This has been confirmed in a large cross-sectional study, where long term sustained weight loss was seen to improve overall metabolic risk (Knell et al., 2018). However, some classically established MetS components, such as HDL cholesterol, are seldom present within the largest cliques, indicating a more peripheral role within this network. Some of the biomarkers we used have a high flow betweenness in the network, suggesting that they behave as an “exchange currency” among several metabolic subsystems. This was the case for triglycerides, insulin, uric acid and glucose, whether considered as physiological parameters or as pathological states (Figure 2). This suggests that they are key components in the transmission of disruptions between different metabolic subsystems. Additionally, these metabolic subsystems, as identified by our clustering strategies, are also those that would be considered as the natural ones from a medical perspective (Chan & Loscalzo, 2012; Goh et al., 2007). Our results show that different, relatively independent, metabolic modules arise, that communicate through some gatekeeping exchange molecules. With age, this modularity increases in the case of the physiological variables network (Figure 3C). Such modularity is a measure of how much the networks tend towards a community structure. Furthermore, there is a strong correspondence between the clusters that were found in the physiological variables network and those found in the pathological states network, suggesting that the associated pathological states emerged from the underlying relationships between the corresponding physiological variables and are, therefore, not just a byproduct of chance or prevalence alone. These two approaches complement each other, reinforcing their respective conclusions where both reach similar results. This was the case for the clustering of metabolic components in both the physiological variables networks, the pathological states networks (Figure 1E and 1F), and the corresponding centrality measurements (Figure 2).

Finally, it is worth mentioning that another advantage of network analysis is that it can be used as part of an automated process for discovering and analyzing patterns in large datasets, with the assistance of experts to ensure a relevant and adequate interpretation (Merico, Gfeller, & Bader, 2009). In this way, networks can be extended in an iterative process in order to accommodate new biomarkers in a way that can both enrich and refine the generated network models (Aittokallio & Schwikowski, 2006). Our work provides the layout for an evidence-based rationale for adding (or replacing) other CVD risk factors (e.g., CRP or family history) to the definition of MetS (Kahn et al., 2005). For instance, the physiological variables network does not rely on the particular values of cut-offs and illustrates that some variables that are not monitored currently, such as uric acid, may be better early indicators of metabolic burden. It is important to notice that uric acid is not used traditionally as a biomarker of metabolic disorders, even when in our network analysis it is more frequently embedded within the largest cliques than blood pressure components, triglycerides and HDL cholesterol (Figure 1C). This result adds to the growing body of literature that considers uric acid to be a relevant biomarker in MetS (Kanbay et al., 2016). In summary, the physiological network approach to metabolic homeostasis is capable of providing useful insights on whole-system function that are inaccessible through reductionist approaches.

### Conclusion

Changes in network topology are global indicators of metabolic homeostasis and do not rely on any single parameter or threshold but, instead, assess the behavior of the whole system. Thus, this novel conceptualization of homeostatic health allows for a more holistic comprehension of a person’s physiology. Structural properties, such as weighted transitivity or the small-world index, may then serve as topological indicators of health for the metabolic physiological network.

## 4. Methodology

### 4.1 Ethical and human research considerations

This study was carried out in accordance with current regulation contained in the Mexican Official Normativity, N0M-012-SSA3-2012. The Ethics Committee of the Facultad de Medicina of the UNAM approved the procedures and protocols for this study under project FM/DI/023/2014, all the participants provided a written informed consent.

### 4.2 Study population and age sub-groups

We performed a transversal, community-based study of an ethnically and educationally diverse sample within a large public university, comprising 2572 participants. Each participant answered a health questionnaire and underwent vital signs, and anthropometric measurements along with fasting blood tests. This resulted in a multi-dimensional data set. The sampling was performed in successive steps from 2014 to 2019. The global sample was divided into 6 groups of increasing age, beginning with less than 25 years, and increasing in decades up to above 65 years of age. As a result, we obtained 6 age groups (see Table 1).

### 4.3 Anthropometric measurements and laboratory procedures

All tests were performed in the morning during a 4-hour period (from 6 a.m. to 10 a.m.) after verifying fasting and general status. Anthropometric measurements (weight, height, waist and hip circumferences) and vital signs (blood pressure and temperature) were taken by trained medical staff using standard procedures (Whelton et al., 2018; WHO, 1995). Blood samples were obtained from participants who had fasted for 8 to 12 hours. Samples were stored at 4-5 °C and submitted for chemical analysis of glucose, glycated hemoglobin (HbA1c), insulin, triglycerides, total cholesterol, HDL cholesterol, LDL cholesterol, uric acid and creatinine. Fasting plasma glucose was measured using spectrophotometry and potentiometry with a hexokinase kit (amorting PIPES, NAD, Hexokinase, ATP, Mg^2^+, G6P-DH; AU 2700 Beckman Coulter R). HbA1c was measured with High Performance Liquid Chromatography (HPLC) analysis with the Variant R Turbo kit 2.0, which consisted of 2 buffers and 1 wash solution. Fasting plasma insulin concentrations were determined using Chemiluminescence (Access Ultrasensitive Insulin, Unicell Dxl 800 Beckman Coulter R, Sensitivity: 0.03-300 U/mL). The lipid profile was obtained with enzymatic colorimetric assay (glycerol phosphate oxidase, cholesterol oxidase, accelerator-selective, detergent, and liquid-selective detergent). Uric acid was measured using the colorimetric method with uricase enzymatic OSR6698, system AU2700/5400, Beckmann Coulter R. This resulted in a set of 15 non-derivative, independent, continuous, physiological variables. From the original data set, 14 particular values associated with distinct variables were excluded, based on two main criteria:

- 1. Outliers based on physiologically improbable values that are most likely to be erroneous as they would be incompatible with life. This included removing three values of blood pressure, three values of axillar temperature, two glucose measurements, two values of HbA1c, and one each of uric acid, and LDL.
- 2. Anthropometric measurements which were inconsistent between themselves. For example, exceedingly high values of waist circumference in an underweight participant. Thirteen values of waist and one value of height were discarded on this account.

### 4.4 Pathological states assessment

From these physiological variables, thresholds were defined in order to distinguish normal values from abnormal values, thus categorizing health status or a pathological state (Table 2). We would like to emphasize that the thresholds used here are not diagnostic of disease, instead they are low enough values that indicate increased risk. Most of our criteria are backed up by major health societies and organizations, however, when a consensus was not available, we used literature-based cut-off values that best correlated with the increased risk-prevention view of the harmonized MetS criteria (Alberti et al., 2009; American Diabetes Association, 2020; Esteghamati et al., 2009; Khanna et al., 2012; Levin et al., 2013; Mach et al., 2019; Sund-Levander, Forsberg, & Wahren, 2002; Tyagi & Aeddula, 2019; Whelton et al., 2018). Thus, the pathological states described here are not diseases per se, but an indication that physiological values do not represent normal health status. Three of the physiological variables that we measured do not have a pathological state by themselves alone. For instance, high blood pressure was determined by either elevated systolic or diastolic values. For insulin and creatinine, two derived indices were calculated: Homeostasis Model Assessment Insulin Resistance index (HOMA-IR) (Wallace, Levy, & Matthews, 2004) for the pathological state of insulin resistance, and eGFR for chronic kidney disease (Levin et al., 2013).

### 4.5 Network modelling

It has been observed that two models of metabolism are possible. In the first one metabolic risk increases progressively as an increasing function of certain physiological variables (Knell et al., 2018; Wijndaele et al., 2006). In the second one, metabolic homeostasis is bimodal, and as such, risk increases significantly only upon exceeding certain thresholds associated with the diagnosis of the pathological state (Alberti et al., 2009; Stern et al., 2005). Therefore, to encompass both possibilities, we created a network model for both employing accessible biomarkers that probe the underlying metabolism.

In the first case, the coupling between two physiological variables can be explored through their rate of change in the population. Here, a monotonic association would be found between those variables that interact directly or indirectly within the physiological network. We tested the physiological variables datasets for normality using the Shapiro-Wilk test and screened them for extreme values. Since the data sets were not normally distributed and had extreme values expected to be real, we selected the Spearman Rank Correlation (Batushansky, Toubiana, & Fait, 2016) as a measure of correlation. We modeled the metabolic physiological network as a continuous association of pairs of variables. For this monotonic correlation model, a correlation matrix was constructed for the 15 chosen physiological parameters (Figure 4). Significant correlations were established at a value of p<0.001, indicating that the relation does not support the null hypothesis that the independent and dependent variables are unrelated. The weight of the Spearman’s rho correlation was squared in order to obtain only positive values.

For the second case, a pathological states network was constructed using currently accepted thresholds from the literature. Here, cut-off values allow the comparison of the tails of the distributions across age groups. The objective here was to indicate whether the participants within the tail of the distribution of one physiological variable have a greater probability of being also in the tail of the distribution of another physiological variable than would be explained by the prevalence of the pathological states alone. This probability of being in a pathological state B given that the individual is in a pathological state A was described using the following binomial test:

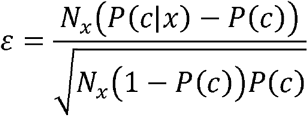

This test is not necessarily reciprocal, thus giving a weighted directionality to the relationship. If a pathological state is probably the origin of another, their value would be expected to be high in that direction, while it could be low in the opposite one. For this binomial test the null hypothesis is that the probability of presenting condition C is not affected by having condition X. The statistical significance, ε, is a measure of the extent to which the null hypothesis is verified by the data. In the circumstance, which is valid here, where the binomial distribution can be approximated by a normal distribution, *ε* > 1.96 corresponds to the standard 95% confidence interval (Easton, Stephens, & Angelova, 2014). As the pathological states network is based upon thresholds accepted by medical consensus, this network adheres well to the known progression of MetS. However, the employment of cut-off values for asserting associations between states may result in an association towards the most sensitive, low thresholds. Exceedingly low thresholds can make pathological states seem more prevalent and bias the direction of £ (Easton et al., 2019). In consequence, care was taken for the selection of thresholds consistent with the preventive scope of MetS.

In summary, for the first case, physiological variables are monotonically correlated along all their biologically plausible spectrum. In this scenario the associations between parameters are present even at healthy values and represent a continuum. For the second case, pathological states are best regarded as binomial. Upon reaching a threshold, the association between these states either appears or increases significantly. This second model resembles the current interpretation of MetS, as it requires a co-occurrence higher than would be expected by chance and contemplates cutoff values as all or nothing states (Alberti et al., 2009). Finally, we used groups of individuals of different ages in order to explore the progressive changes that occur during the aging process and which result in an increasing prevalence of MetS. From the systems biology perspective, the network structure is a direct result of the coordination, or lack thereof, of components that are linked by homeostatic feedback (Goldstein, 2019).

### 4.6 Network construction and statistical analysis

For the construction of our considered networks we used correlation matrices of physiological variables and pathological states. These matrices were interpreted as weighted adjacency matrices, where adjacency is represented by the Spearman rhos or the values between each pair of metabolic components. The resulting matrices were weighted and undirected for the Pearson correlation matrix and weighted and directed in the case of ε values (Figure 4). For the construction of the Spearman correlation matrix, data-set normality testing, linear regression and chi-squared tests for trends were all done with Prism 8.1.2(277), GraphPad Software, La Jolla, California USA, www.graphpad.com. For the network construction RStudio, an R language programing suite and igraph package (Csárdi, Nepusz, & Airoldi, 2016; R Core Team, 2020; RStudio Team, 2020).

Nodes within a network can be ranked according to several centrality definitions that fall into two main groups, radial measures and medial measures. Inferring causality exclusively from centrality within networks, requires caution, although eigencentrality has been found to be the best centrality measurement for this purpose, especially for small networks with less than 30 nodes (Dablander & Hinne, 2019). Therefore, we selected eigenvector for undirected networks and hubscore for directed networks as radial measures. For medial measures we decided to use flow betweenness. These centrality values allow for a direct comparison of either the influence of nodes (radial measure) or gatekeeping (medial measure) within the network (Borgatti & Everett, 2006). Eigencentrality corresponds to the value of the first eigenvector of the graph adjacency matrix and was interpreted as a measure of influence within the undirected networks. These values were obtained using the evcent function from the SNA package (Butts, 2019). For directed networks, hub score and authority score, are a better way of representing influence as these measures take into account the directionality of the links. Hub scores are defined as the principal eigenvectors of A*t(A), where A is the adjacency matrix of the network. These values were calculated with the hub_score function from the igraph package (Kleinberg, 1998). Flow betweenness was used as a measurement of intermediation within the network. Flow betweenness was calculated using the flowbet function from the SNA package. In order to test if the eigencentrality and flow betweenness values obtained would be seen in a random graph with the same number of vertices, edges or dyads, univariate conditionally uniform graph tests (CUG test) were employed with the cug.test function from the SNA package.

Networks can contain subgraphs, subsets of vertices with a specific set of edges connecting them within the original graph, that are of particular relevance (Aittokallio & Schwikowski, 2006). We sought two particular subgraphs within our models: First, the graph corresponding to those variables associated with the current definition of MetS, and second, the largest clique within the graph. As there may be more than one combination of nodes that result in a largest clique, we registered the number of times each node appeared within a possible largest clique. These maximally connected subgraphs - largest cliques - were identified using the largest_cliques function of the igraph package (Eppstein, Löffler, & Strash, 2010). Largest clique and current MetS variables were highlighted as subgraphs, along with the graph diameter.

The largest clique is the biggest, maximally connected subgraph of a graph and contains vertices such that each vertex is connected with every other vertex of the clique. This gives an idea of which vertices go hand in hand in each network (Pavlopoulos et al., 2011). On the other hand, a cluster, as defined using a suitable clustering algorithm, is a group of vertices within a graph that are more densely connected to one another than to other vertices (Csárdi et al., 2016). There are several alternative algorithms for discovering communities of vertices within graphs. For community detection within the networks we used two different algorithms. For the Pearson model, the Louvain algorithm was employed as a heuristic method based on modularity optimization, with the cluster_louvain function from the igraph package (Blondel et al., 2008). In the £ model, the spinglass community algorithm selects those nodes with the greatest probability to be found in the same state concurrently, with the cluster_spinglass function from the igraph package (Reichardt & Bornholdt, 2006). These two approaches to identifying related biomarkers are complementary - clustering strategies maximize the modularity of the network, while largest-clique identification maximizes the transitivity of the largest possible subgraph.

Topological properties were assessed as follows: Density, reciprocity and characteristic path length of the networks were calculated using the igraph package. For the calculation of the weighted transitivity and the clustering coefficient in directed and undirected weighted networks the DirectedClustering package was employed (Clemente & Grassi, 2018). The Small world index, as calculated by qgraph, was used as a summary metric of the network topology (Watts & Strogatz, 1998). CUG tests were also performed for network density, reciprocity, transitivity and characteristic path length.

## Data Availability

The datasets associated with this study can be found publicly available in either csv or database formats at [http://project42.c3.unam.mx/].

## 6 Author Contributions

CRS designed the project and obtained the funding. ABM conceived this work, implemented the network modelling and with JFE and ALR performed all the statistical and network analysis. ABM, RMT, and LC contributed with the acquisition and the medical interpretation of the data. All authors contributed with the manuscript revision, read and approved the submitted version.

## 8 Acknowledgments

Antonio Barajas is a doctoral student from Programa de Doctorado en Ciencias Biomédicas, Universidad Nacional Autónoma de México (UNAM) and received fellowship 596756 from CONACYT. We thank the Instituto Nacional de Ciencias Médicas y Nutrición “Salvador Zubirán” and the Hospital Juárez de México for the sample processing described in the laboratory procedures.

## References

Aittokallio, T., & Schwikowski, B. (2006). Graph-based methods for analysing networks in cell biology. Briefings in Bioinformatics, 7(3), 243–255. https://doi.org/10.1093/bib/bbl022

Alberti, K. G. M. M., Eckel, R. H., Grundy, S. M., Zimmet, P. Z., Cleeman, J. I., Donato, K. A., … Smith, S. C. (2009). Harmonizing the metabolic syndrome: A joint interim statement of the international diabetes federation task force on epidemiology and prevention; National heart, lung, and blood institute; American heart association; World heart federation; International. Circulation, 120(16), 1640–1645. https://doi.org/10.1161/CIRCULATIONAHA.109.192644

Almeda-Valdes, P., Aguilar-Salinas, C. A., Uribe, M., Canizales-Quinteros, S., & Méndez-Sánchez, N. (2016). Impact of anthropometric cut-off values in determining the prevalence of metabolic alterations. European Journal of Clinical Investigation, 46(11), 940–946. https://doi.org/10.1111/eci.12672

American Diabetes Association. (2020, January 1). 2. Classification and Diagnosis of Diabetes: Standards of Medical Care in Diabetes-2020. Diabetes Care. NLM (Medline). https://doi.org/10.2337/dc20-S002

Barabási, A.-L., Dezső, Z., Ravasz, E., Yook, S., & Oltvai, Z. (2003). Scale-Free and Hierarchical Structures in Complex Networks. In AIP Conference Proceedings (Vol. 661, pp. 1–16). AIP Publishing. https://doi.org/10.1063/1.1571285

Batushansky, A., Toubiana, D., & Fait, A. (2016). Correlation-Based Network Generation, Visualization, and Analysis as a Powerful Tool in Biological Studies: A Case Study in Cancer Cell Metabolism. BioMed Research International, 2016. https://doi.org/10.1155/2016/8313272

Blondel, V. D., Guillaume, J.-L., Lambiotte, R., & Lefebvre, E. (2008). Fast unfolding of communities in large networks. Journal of Statistical Mechanics: Theory and Experiment, 2008(10), P10008.

Borgatti, S. P., & Everett, M. G. (2006). A Graph-theoretic perspective on centrality. Social Networks, 28(4), 466–484. https://doi.org/10.1016/J.SOCNET.2005.11.005

Broido, A. D., & Clauset, A. (2019). Scale-free networks are rare. Nature Communications, 10(1), 1–10. https://doi.org/10.1038/s41467-019-08746-5

Butts, C. T. (2019). sna: Tools for Social Network Analysis. https://cran.r-project.org/package=sna

Chan, S. Y., & Loscalzo, J. (2012, July 20). The emerging paradigm of network medicine in the study of human disease. Circulation Research. Lippincott Williams & WilkinsHagerstown, MD. https://doi.org/10.1161/CIRCRESAHA.111.258541

Chen, P., Li, Y., Liu, X., Liu, R., & Chen, L. (2017). Detecting the tipping points in a three-state model of complex diseases by temporal differential networks. Journal of Translational Medicine, 15(1), 217. https://doi.org/10.1186/s12967-017-1320-7

Csárdi, G., Nepusz, T., & Airoldi, E. M. (2016). Statistical Network Analysis with igraph. Springer. https://sites.fas.harvard.edu/~airoldi/pub/books/BookDraft-CsardiNepuszAiroldi2016.pdf

Dablander, F., & Hinne, M. (2019). Node centrality measures are a poor substitute for causal inference. Scientific Reports, 9(1), 6846. https://doi.org/10.1038/s41598-019-43033-9

Easton, J. F., Robles-Cabrera, A., Fossion, R., Rivera, A. L., & Stephens, C. R. (2019). Thoughts on the use of standard cut-off values for physiological health indicators. AIP Conference Proceedings, 2090, 50006. https://doi.org/10.1063/1.5095921

Easton, J. F., Stephens, C. R., & Angelova, M. (2014). Risk factors and prediction of very short term versus short/intermediate term post-stroke mortality: A data mining approach. Computers in Biology and Medicine, 54, 199–210. https://doi.org/10.1016/J.COMPBIOMED.2014.09.003

Enzi, G., Busetto, L., Inelmen, E. M., Coin, A., & Sergi, G. (2003). Historical perspective: visceral obesity and related comorbidity in Joannes Baptista Morgagni’s ‘De Sedibus et Causis Morborum per Anatomen Indagata.’ International Journal of Obesity, 27(4), 534–535. https://doi.org/10.1038/sj.ijo.0802268

Eppstein, D., Löffler, M., & Strash, D. (2010). Listing All Maximal Cliques in Sparse Graphs in Near-optimal Time. Algorithms and Computation, 6506, 403–414. http://arxiv.org/abs/1006.5440

Esteghamati, A., Ashraf, H., Esteghamati, A.-R., Meysamie, A., Khalilzadeh, O., Nakhjavani, M., & Abbasi, M. (2009). Optimal threshold of homeostasis model assessment for insulin resistance in an Iranian population: the implication of metabolic syndrome to detect insulin resistance. Diabetes Research and Clinical Practice, 84(3), 279–287. https://doi.org/10.1016/j.diabres.2009.03.005

Fossion, R., Rivera, A. L., & Estañol, B. (2018). A physicist’s view of homeostasis: how time series of continuous monitoring reflect the function of physiological variables in regulatory mechanisms. https://doi.org/10.1088/1361-6579/aad8db

Goh, K.-I., Cusick, M. E., Valle, D., Childs, B., Vidal, M., & Szló Barabá, A.-L. (2007). The human disease network. www.pnas.org/cgi/content/full/

Goldstein, D. S. (2019). How does homeostasis happen? Integrative physiological, systems biological, and evolutionary perspectives. American Journal of Physiology. Regulatory, Integrative and Comparative Physiology, 316(4), R301–R317. https://doi.org/10.1152/ajpregu.00396.2018

Haring, R., Rosvall, M., Völker, U., Völzke, H., Kroemer, H., Nauck, M., & Wallaschofski, H. (2012). A Network-Based Approach to Visualize Prevalence and Progression of Metabolic Syndrome Components. PLoS ONE, 7(6), e39461. https://doi.org/10.1371/journal.pone.0039461

Hildrum, B., Mykletun, A., Hole, T., Midthjell, K., & Dahl, A. A. (2007). Age-specific prevalence of the metabolic syndrome defined by the International Diabetes Federation and the National Cholesterol Education Program: the Norwegian HUNT 2 study. BMC Public Health, 7(1), 220. https://doi.org/10.1186/1471-2458-7-220

Hilgetag, C. C., & Goulas, A. (2016). Is the brain really a small-world network? Brain Structure and Function, 221(4), 2361–2366. https://doi.org/10.1007/s00429-015-1035-6

Huang, P. L. (2009). A comprehensive definition for metabolic syndrome. Disease Models & Mechanisms, 2(5–6), 231–237. https://doi.org/10.1242/dmm.001180

Kahn, R. (2007). Metabolic syndrome: is it a syndrome? Does it matter? Circulation, 115(13), 1806–1810; discussion 1811. https://doi.org/10.1161/CIRCULATIONAHA.106.658336

Kahn, R., Buse, J., Ferrannini, E., Stern, M., American Diabetes Association, & European Association for the Study of Diabetes. (2005). The metabolic syndrome: time for a critical appraisal: joint statement from the American Diabetes Association and the European Association for the Study of Diabetes. Diabetes Care, 28(9), 2289–2304. https://doi.org/10.2337/DIACARE.28.9.2289

Kanbay, M., Jensen, T., Solak, Y., Le, M., Roncal-Jimenez, C., Rivard, C., … Johnson, R. J. (2016). Uric acid in metabolic syndrome: From an innocent bystander to a central player. European Journal of Internal Medicine, 29, 3–8. http://www.ncbi.nlm.nih.gov/pubmed/26703429

Khanna, D., Fitzgerald, J. D., Khanna, P. P., Bae, S., Singh, M. K., Neogi, T., … Terkeltaub, R. (2012). 2012 American College of Rheumatology Guidelines for Management of Gout. Part 1: Systematic Nonpharmacologic and Pharmacologic Therapeutic Approaches to Hyperuricemia. https://doi.org/10.1002/acr.21772

Kitano, H., Oda, K., Kimura, T., Matsuoka, Y., Csete, M., Doyle, J., & Muramatsu, M. (2004). Metabolic Syndrome and Robustness Tradeoffs. Diabetes, 53(suppl 3), S6–S15. https://doi.org/10.2337/DIABETES.53.SUPPL_3.S6

Knell, G., Li, Q., Pettee Gabriel, K., & Shuval, K. (2018). Long-Term Weight Loss and Metabolic Health in Adults Concerned With Maintaining or Losing Weight: Findings From NHANES. Mayo Clinic Proceedings, 93(11), 1611–1616. https://doi.org/10.1016/j.mayocp.2018.04.018

Leatherdale, S. T. (2015). An examination of the co-occurrence of modifiable risk factors associated with chronic disease among youth in the COMPASS study. Cancer Causes & Control, 26(4), 519–528. https://doi.org/10.1007/s10552-015-0529-0

Leventhal, A. M., Huh, J., & Dunton, G. F. (2014). Clustering of modifiable biobehavioral risk factors for chronic disease in US adults: a latent class analysis. Perspectives in Public Health, 134(6), 331–338. https://doi.org/10.1177/1757913913495780

Levin, A., Stevens, P. E., Bilous, R. W., Coresh, J., De Francisco, A. L. M., De Jong, P. E., … Winearls, C. G. (2013). Kidney disease: Improving global outcomes (KDIGO) CKD work group. KDIGO 2012 clinical practice guideline for the evaluation and management of chronic kidney disease. Kidney International Supplements. Nature Publishing Group. https://doi.org/10.1038/kisup.2012.73

Lusis, A. J., Attie, A. D., & Reue, K. (2008). Metabolic syndrome: from epidemiology to systems biology. Nature Reviews Genetics, 9(11), 819–830. https://doi.org/10.1038/nrg2468

Mach, F., Baigent, C., Catapano, A. L., Koskinas, K. C., Casula, M., Badimon, L., … Patel, R. S. (2019). 2019 ESC/EAS Guidelines for the management of dyslipidaemias: lipid modification to reduce cardiovascular risk. European Heart Journal. https://doi.org/10.1093/eurheartj/ehz455

Merico, D., Gfeller, D., & Bader, G. D. (2009). How to visually interpret biological data using networks. Nature Biotechnology, 27(10), 921–924. https://doi.org/10.1038/nbt.1567

O’Neill, S., & O’Driscoll, L. (2015). Metabolic syndrome: a closer look at the growing epidemic and its associated pathologies. Obesity Reviews, 16(1), 1–12. https://doi.org/10.1111/obr.12229

Parikh, R., & Mohan, V. (2012). Changing definitions of metabolic syndrome. Indian Journal of Endocrinology and Metabolism, 16(1), 7. https://doi.org/10.4103/2230-8210.91175

Pavlopoulos, G. A., Secrier, M., Moschopoulos, C. N., Soldatos, T. G., Kossida, S., Aerts, J., … Bagos, P. G. (2011). Using graph theory to analyze biological networks. BioData Mining, 4(1), 10. https://doi.org/10.1186/1756-0381-4-10

R Core Team. (2020). R: A language and environment for statistical computing. Viena, Austria: R Foundation for Statistical Computing. https://www.r-project.org/

Reaven, G. M. (1993). Role of Insulin Resistance in Human Disease (Syndrome X): An Expanded Definition. Annual Review of Medicine, 44(1), 121–131. https://doi.org/10.1146/annurev.me.44.020193.001005

Reichardt, J., & Bornholdt, S. (2006). Statistical Mechanics of Community Detection. Physical Review E, Vol. 74, Issue 1, Id. 016110, 74(1). https://doi.org/10.1103/PhysRevE.74.016110

RStudio Team. (2020). RStudio: Integrated Development for R. PBC. Boston, MA: RStudio. http://www.rstudio.com/.

Sattar, N. (2008). Why metabolic syndrome criteria have not made prime time: a view from the clinic. International Journal of Obesity, 32(S2), S30–S34. https://doi.org/10.1038/ijo.2008.33

Song, C., Havlin, S., & Makse, H. A. (2005). Self-similarity of complex networks. Nature, 433(7024), 392–395. https://doi.org/10.1038/nature03248

Stephens, C. R., Easton, J. F., Robles-Cabrera, A., Fossion, R. Y. M., De La Cruz, L., Martinez-Tapia, R., … Rivera, A. L. (2020). The impact of education and age on metabolic disorders. Frontiers in Public Health, 8, 180. https://doi.org/10.3389/FPUBH.2020.00180

Stern, S. E., Williams, K., Ferrannini, E., DeFronzo, R. A., Bogardus, C., & Stern, M. P. (2005). Identification of individuals with insulin resistance using routine clinical measurements. Diabetes, 54(2), 333–339. https://doi.org/DOI:10.2337/diabetes.54.2.333

Sun, L., Yu, Y., Huang, T., An, P., Yu, D., Yu, Z., … Wang, F. (2012). Associations between Ionomic Profile and Metabolic Abnormalities in Human Population. PLoS ONE, 7(6), e38845. https://doi.org/10.1371/journal.pone.0038845

Sund-Levander, M., Forsberg, C., & Wahren, L. K. (2002). Normal oral, rectal, tympanic and axillary body temperature in adult men and women: a systematic literature review. Scandinavian Journal of Caring Sciences, 16(2), 122–128. https://doi.org/10.1046/j.1471-6712.2002.00069.x

Toledo-Roy, J. C., Rivera, A. L., & Frank, A. (2019). Symmetry, criticality and complex systems. In Symmetries and Order: Algebraic Methods in Many Body Systems: A symposium in celebration of the career of Professor Francesco Iachello (Vol. 2150, p. 020014). AIP Publishing. https://doi.org/10.1063/1.5124586

Tyagi, A., & Aeddula, N. R. (2019). Azotemia. StatPearls Publishing, Treasure Island (FL). http://europepmc.org/books/NBK538145

Vassallo, P., Driver, S. L., & Stone, N. J. (2016). Metabolic Syndrome: An Evolving Clinical Construct. Progress in Cardiovascular Diseases, 59(2), 172–177. https://doi.org/10.1016/j.pcad.2016.07.012

Vona, R., Gambardella, L., Cittadini, C., Straface, E., Pietraforte, D., Editor, G., & Di Mauro, M. (2019). Biomarkers of Oxidative Stress in Metabolic Syndrome and Associated Diseases. Oxidative Medicine and Cellular Longevity, 2019, 1–19. https://doi.org/10.1155/2019/8267234

Wallace, T. M., Levy, J. C., & Matthews, D. R. (2004). Use and Abuse of HOMA Modeling. Diabetes Care, 27(6), 1487 LP – 1495. https://doi.org/10.2337/diacare.27.6.1487

Whelton, P. K., Carey, R. M., Aronow, W. S., Casey, D. E., Collins, K. J., Dennison Himmelfarb, C., … Wright, J. T. (2018). 2017 ACC / AHA / AAPA / ABC / ACPM / AGS / APhA / ASH / ASPC / NMA / PCNA Guideline for the Prevention, Detection, Evaluation, and Management of High Blood Pressure in Adults: Executive Summary. Journal of the American College of Cardiology, 71(19), 2199–2269. https://doi.org/10.1016/j.jacc.2017.11.005

WHO. (1995). Physical status: the use and interpretation of anthropometry. Report of a WHO Expert Committee. World Health Organization technical report series (Vol. 854). Switzerland.

Wijndaele, K., Beunen, G., Duvigneaud, N., Matton, L., Duquet, W., Thomis, M., … Philippaerts, R. M. (2006). A continuous metabolic syndrome risk score: Utility for epidemiological analyses [6]. Diabetes Care, 29(10), 2329. https://doi.org/10.2337/dc06-1341

Xu, H., Li, X., Adams, H., Kubena, K., & Guo, S. (2018). Etiology of Metabolic Syndrome and Dietary Intervention. International Journal of Molecular Sciences, 20(1), 128. https://doi.org/10.3390/ijms20010128

